# Comparison of procedures performed and costs between an outsourced dental health plan with fee-for-service payment and an evidence-based, in-house dental health service with salaried professionals

**DOI:** 10.1101/2022.03.27.22273012

**Authors:** Luiz Cesar Da Costa Filho, Carolina Covolo Da Costa, Carisi Anne Polanczyk, Bruce Bartholow Duncan

## Abstract

It was evaluated a dental care services provided to almost 4,000 users (employees and their families) of a company (Moinhos de Vento Hospital, Brazil). The analysis was divided into two periods of time: from January 2000 to December 2002, during which time the dental care provided to employees (and families) was outsourced to a dental plan which operates with an accredited network, and from March 2003 to June 2005, when the dental care was provided by an in-house dental care service with preventive and evidence-based dentistry guidelines. Economic data were collected, as well as the type and number of procedures performed.

**Conclusions:** An in-house service with evidence-based clinical protocols, as compared with an accredited network, decreased costs and also increased the number of procedures done, changed the profile of care for a less invasive dentistry and one more geared toward the causes of diseases, while still preserving the customers’ satisfaction.

## INTRODUCTION

In the last few decades, low and middle income countries have witnessed an increased number of dentists and the continuity of problems related to dental care services access for less-privileged members of society. That is Brazil’s reality, where dental care assistance is going through a paradoxical stage, because Brazil is one of the countries with the highest number of dentists worldwide (371,112; 1 dentist per 578 inhabitants) (1) and at the same time more than 10% of its population has never had access to any type of dental care assistance (2). Therefore, dental care assistance offered by companies to their employees is highly relevant because it allows access to oral health services, in addition to offering to new dental professionals their entry into the workforce (3). The dental care medicine sector is the fastest-growing segment of benefits provided by Brazilian companies (4, 5). At the same time, however, the increased costs of employee health are a cause of concern for many companies. (6-8) Within this context, evidence-based health practices could assist businesses in expanding health benefits, because if they adopt clinical procedures with more efficiency, it will optimize their financial resources and amplify the quality of health care (9-14). This article aims to retrospectively assess the costs, coverage, care profile, and customer satisfaction in relation to dental services offered by the “Fundação de Amparo Social do Hospital Moinhos de Vento (FAS-HMV)” foundation. This foundation aims to offer a health plan for employees (and their families) of the Moinhos de Vento Hospital (HMV), which is a private hospital with 360 hospital beds located in Porto Alegre, RS, Brazil. The evaluation was made at two points: before the implementation of an in-house dental service—a period during which an outsourced dental health plan with an accredited network provided dental care procedures—and after the implementation of an in-house evidence-based service.

## METHODS

### The dental service of FAS-HMV foundation

The FAS-HMV foundation was created in 1995 with a main objective of implementing a self-managed health plan for employees at the Moinhos de Vento Hospital (HMV) and related entities. The plan also benefits employees’ spouses and children (until the age of 21 years old), retired employees, and ex-employees according to Brazilian law 9656/98. In 1995, when the health plan of the FAS-HMV foundation was established, dental care was outsourced to a company that provides dental care services and operates with a network of accredited professionals, who were remunerated through a fee-for-service system. In mid 2002 another dental care company was hired to do an epidemiological survey for the population of users from the FAS-HMV foundation and thereby estimate and verify the possibility of implementing an in-house dental service. In February 2003, along with other structural changes in the FAS-HMV foundation health plan, the in-house service for dental care was implemented. The sizing of the service and the clinical profile of professionals were planned based on information from the 2002 epidemiological survey. The proposal of service for dental care aimed to promote oral health, which aside from providing for the clinical needs of the patient, emphasized prevention and oral health education. Another characteristic of this service is that clinical protocols were elaborated with a methodology of evidence-based health practices, aiming to optimize the economic resources related to the self-managed health plan.

During the period between 2003 and 2005, the in-house service worked with 3 dental offices, 7 dentists, 1 dental hygiene technician, and 2 dental assistants. The profile of the selected dentists for the service was one of general clinicians with high-quality training, but who also had different specialties (dentistry, pediatric dentistry, orthodontics, endodontics, periodontics, and oral surgery).

In order to increase the resolution rate of complex cases at the in-house service, the team of professionals chosen also had knowledge of evidence-based dentistry concepts so they could also contribute to the elaboration of clinical protocols. The dental hygiene technician and the dental office assistants had an oriented training to perform basic procedures in oral health (supragingival scraping and oral health education).

Unlike the previous system, remuneration was based on salaries rather than a fee-for-services payment system.

### Design

This paper reports an Interrupted Time Series assessed at two different moments in the company’s history, based on secondary data collected at the FAS-HMV foundation from January 2000 to June 2005. This research was approved by the ethics committee and the scientific committee from the Education and Research Institute of the Moinhos de Vento Hospital (Project CEP IEP-HMV: #2005/38).

### Study population and sample size

The study population was users of the FAS-HMV foundation health plan. The foundation’s health plan had almost 4,000 users, this being an ever-changing number according to inflow and outflow of employees and their families. Users were employees (active, retired and ex-employees) from Moinhos de Vento Hospital and related entities, with or without their spouses and children up to 21 years of age.

### Methods

Information related to procedures performed in 2000, 2001 and 2002 was provided by the outsourced dental plan company responsible for dental care. They also provided customer satisfaction surveys from the years 2000 and 2001.

Data from March 2003 to 2005 were obtained in the clinical records of the FAS-HMV foundation in-house dental care service. The survey of service satisfaction during this period of time was obtained by the foundation through a series of monthly questionnaires in randomally sampled service users. Data about dental care costs from January 2000 to June 2005 was obtained from the company responsible for the management of the health plan from the FAS-HMV foundation since its creation in 1995. Costs measured before the in-house service included: *I*) the amount paid to the outsourced dental plan; *II*) dental emergencies expenses; *III*) costs of specialized care, outside the operators network; *IV*) Costs of the hospital’s surgical services, hospitalizations, and exams; and *V*) related salaries and charges generated by employee absences due to dental reasons. With reference to the in-house service, the following costs were taken into account: *I*) costs of salaries and charges of dental health staff; *II*) costs related to materials purchasing and equipment maintenance; *III*) expenses related to dental emergencies; *IV*) costs of specialized care outside the in-house service; *V*) costs of the hospital’s surgical services, hospitalizations and exams; *VI*) expenses related to general services (water, light, telephone, rent, cleansing, etc.); *VII*) Costs related to equipment depreciation; *VIII*) Costs related to possible labor liabilities, and *IX*) related salaries and charges generated by employee absences due to dental reasons.

### Statistical analysis

Data concerning dental care procedures and costs were processed using the statistical software package SPSS 10.0.1 and Microsoft Office Excel 2003. Data information was collected monthly during the 36 months prior to the changes in the dental service system and during the 28 months following. Considering the variation in the number of beneficiary employees of the program and aiming to allow a greater comparability between the two periods of time, results were standardized for 4000 users. Expenses were adjusted monthly by Brazilian IPCA inflation index (Broad Consumers Price Index: monitored prices—health plans) and then were converted into International Dollars for the year 2003 according to tables of purchasing power parity by the World Health Organization (WHO) and Organization for Economic Co-operation and Development (OECD) (12,15). The historical series of the IPCA were obtained through the website of the Brazilian Central Bank (serie # 4461) (16).

Comparisons between the two dental service management periods—the outsourced dental plan (with an accredited network of dentists) and the in-house service—were done using a t-test for independent samples or a Mann-Whitney’s test, depending on data distribution. The significance level of 5% was established for statistical differences.

The time-trend graphs were plotted using Lowess function with 3 interactions and using 50% of the points for adjustment.

## RESULTS

In the diagnostic area (table 1), no quantitative difference exists in the total number of procedures between the systems, but there is a difference regarding the type of procedures. Whereas the accredited network had a greater number of clinical examinations, the in-house service had a greater number of radiographs. These might be attributed to the fact that professionals from the accredited network generated more clinical examinations, whereas the professionals from in-house service had access to the same dental records with an initial dental planning, which could be shared by other professionals in treatment consultations between different experts of the same service.

**Table 1:** Number of procedures of the most prevalent areas (diagnostics, prevention, oral environment adequacy, restorative) – Monthly means and standard deviation.

| Area | Procedures | Outsourced<br>Dental Plan | In-house Dental<br>Service |
| --- | --- | --- | --- |
|  |  | N ± SD | N ± SD |
| Diagnostics | Initial exam and dental planning | 155 ± 88 | 116 ± 75 |
|  | Clinical appointments for examination ** | 129 ± 72 | 56 ± 15 |
|  | Radiographs ** | 131 ± 48 | 236 ± 54 |
|  | Biopsies ** | 0,0 ± 0,0 | 0,4 ± 0,8 |
|  | **Total - Diagnostics** | **415 ± 116** | **409 ± 43** |
| Prevention | Prophylaxis or Oral Hygiene Instructions ** | 192 ± 62 | 298 ± 65 |
|  | Fluoride Therapy ** | 77 ± 47 | 208 ± 43 |
|  | Dental Sealant per Tooth ** | 2 ± 6 | 14 ± 9 |
|  | **Total – Prevention \*\*** | **271 ± 103** | **520 ± 104** |
| Oral<br>Adequacy<br>Procedures | Supra and sub gingival scaling and root<br>planning** | 48 ± 24 | 228 ± 57 |
|  | Glass Ionomer restorations ** | 5 ± 5 | 29 ± 11 |
|  | Restorations reconditioning ** | 8 ± 6 | 20 ± 9 |
|  | Expectant treatments ** | 6 ± 5 | 38 ± 10 |
|  | **Total – Oral Adequacy Procedures \*\*** | **67 ± 32** | **315 ± 74** |
| Restorative | Tooth Fragment Bonding | 0,1 ± 0,3 | 0,1 ± 0,4 |
|  | Reconstructions (amalgam or composite) ** | 5 ± 4 | 1 ± 1 |
|  | Amalgam Restorations ** | 50 ± 35 | 21 ± 11 |
|  | Composite Restorations ** | 242 ± 77 | 87 ± 23 |
|  | **Total – Restorative \*\*** | **297 ± 99** | **109 ± 25** |

Statistical differences: \* p<0,05 \*\*p<0,01.

With the in-house service, we observed an increase in the number of more conservative procedures, which are preventive actions (table 1) and oral environment adequacy actions, which include basic periodontics procedures, provisional restorations, glass ionomer restorations, expectant treatments, and restorations repairs (repolishing, excess removal, and small restoration repairs) (table 1). This increase could be a reflection of evidence-based clinical protocols, which are strongly related to the treatment of the causes of the underlying pathologies of the problems which presented (17-23) and not merely treatment of symptoms. Among the restorative procedures (table 1), there was a sharp reduction with the in-house service. This could be explained by the possible existence within the accredited network of a professional tendency to change restorations with the slightest evidence of need, or because within the in-house service a greater number of reconditioning and repairs of restorations was observed. Another potential explanation is that within the in-house service, when no need for immediate treatment existed, the case can be more closely observed prior to undertaking a therapeutic intervention, which may have resulted in more radiographic observations over time (table 1).

The in-house service also brought a decline in dental extractions (table 2), which could be an indicator of conservative protocols for the extraction of teeth with cavities or periodontal disease, as well as a more conservative protocol for the removal of impacted teeth (24).

**Table 2:** Tooth extractions and endodontic treatments – Monthly means and standard deviation.

| Area | Procedures | Outsourced<br>Dental Plan | In-house Dental<br>Service |
| --- | --- | --- | --- |
|  |  | N ± SD | N ± SD |
| Tooth<br>extraction | Tooth extraction ** | 24 ± 11 | 16 ± 8 |
|  | Impacted tooth extraction * | 12 ± 6 | 8 ± 4 |
|  | Deciduous tooth extraction ** | 8 ± 8 | 12 ± 5 |
| Endodontics | Direct pulp capping | 3 ± 3 | 2 ± 2 |
|  | Endodontic treatment * | 16 ± 7 | 12 ± 6 |
|  | Endodontic re-treatment ** | 3 ± 3 | 1 ± 1 |
|  | Pulpotomy | 1 ± 1 | 1 ± 1 |
|  | Deciduous endodontic treatment | 0,3 ± 0,5 | 0,9 ± 1,2 |
|  | Deciduous Pulpotomy | 0,2 ± 0,6 | 0,1 ± 0,4 |

Statistical differences: \* p<0,05 \*\*p<0,01.

The number of root canal treatments also decreased with the adoption of the in-house service (table 2). This may have occurred because the increase of temporary restorations with glass ionomer (table 1), which allowed the softened dentine to remineralize, and resulted in fewer interventions on dental pulp in the restorative stage. Additionally, decreases in restoration changes may also have resulted in fewer pulp invasions. Another, less likely explanation, due to the short amount of time covered in the service evaluation, is that the increase in preventive actions resulted in a lower incidence of caries.

The decrease of endodontic retreatment by the in-house service could be explained through more observational and follow-up protocols, actions that may have also contributed to the increase in the number of radiographs (table 1).

In terms of other low-demand odontopediatric services, we found that the in-house service conducted more dental extractions of deciduous teeth. With respect to the number of pulpotomies and root canals in primary teeth, we found no difference between the two phases (table 2).

The in-house service increased in two new areas: occlusion, and preventive and interceptative orthodontics (table 3). Is important to mention that the dental plan users only paid the laboratory costs of the dental appliances.

**Table 3:** Minor oral surgery, occlusion, orthodontics, and other procedures - Monthly means and standard deviation.

| Area | Procedures | Outsourced<br>Dental Plan | In-house Dental<br>Service |
| --- | --- | --- | --- |
|  |  | N ± SD | N ± SD |
| Minor Oral Surgery | Gingivectomy ** | 2,9 ± 2,7 | 1,0 ± 1,0 |
|  | Advanced Periodontal Surgery ** | 0,1 ± 0,3 | 5,9 ± 3,7 |
|  | Other minor oral surgery * | 0,0 ± 0,0 | 0,1 ± 0,3 |
| Occlusion | Occlusal adjustments** | 0,1 ± 0,4 | 5 ± 3 |
|  | Oral Appliances ** | -- | 4 ± 3 |
| Orthodontics | Orthodontic appointment ** | -- | 33 ± 9 |
|  | Orthodontic Appliances ** | -- | 4 ± 2 |
| Other Procedures | Emergency appointment ** | 18 ± 7 | 34 ± 9 |
|  | External Specialist Appointment<br>** | 5 ± 3 | 1 ± 2 |
|  | Hospital surgical appointment * | 0,6 ± 0,8 | 0,3 ± 0,5 |

Statistical differences: \* p<0,05 \*\*p<0,01.
-- not covered procedure

In table 3, the results suggest that minor surgeries increased with the in-house service, and simpler surgeries like gingivectomy were replaced by more complex periodontal procedures such as clinical crown lengthening, access flaps and regenerative procedures.

In relation to specific services (table 3), the results show that the in-house service performed more emergency treatments, with less use of experts’ services outside their own professional staff, and fewer hospital services. However, there may be a underrecording of emergencies within the accredited network, due to the fact that professionals use different coding patterns within the two health systems.

The in-house dental service performed a higher quantity of procedures with lower costs (table 4). Nevertheless, the procedures profile was different between the two services: whereas in the accredited network dental plan, diagnostic and restorative procedures predominated, in the in-house service diagnostic services, prevention, and adequacy of oral environment procedures were most frequent (Figure 1).

**Table 4:**
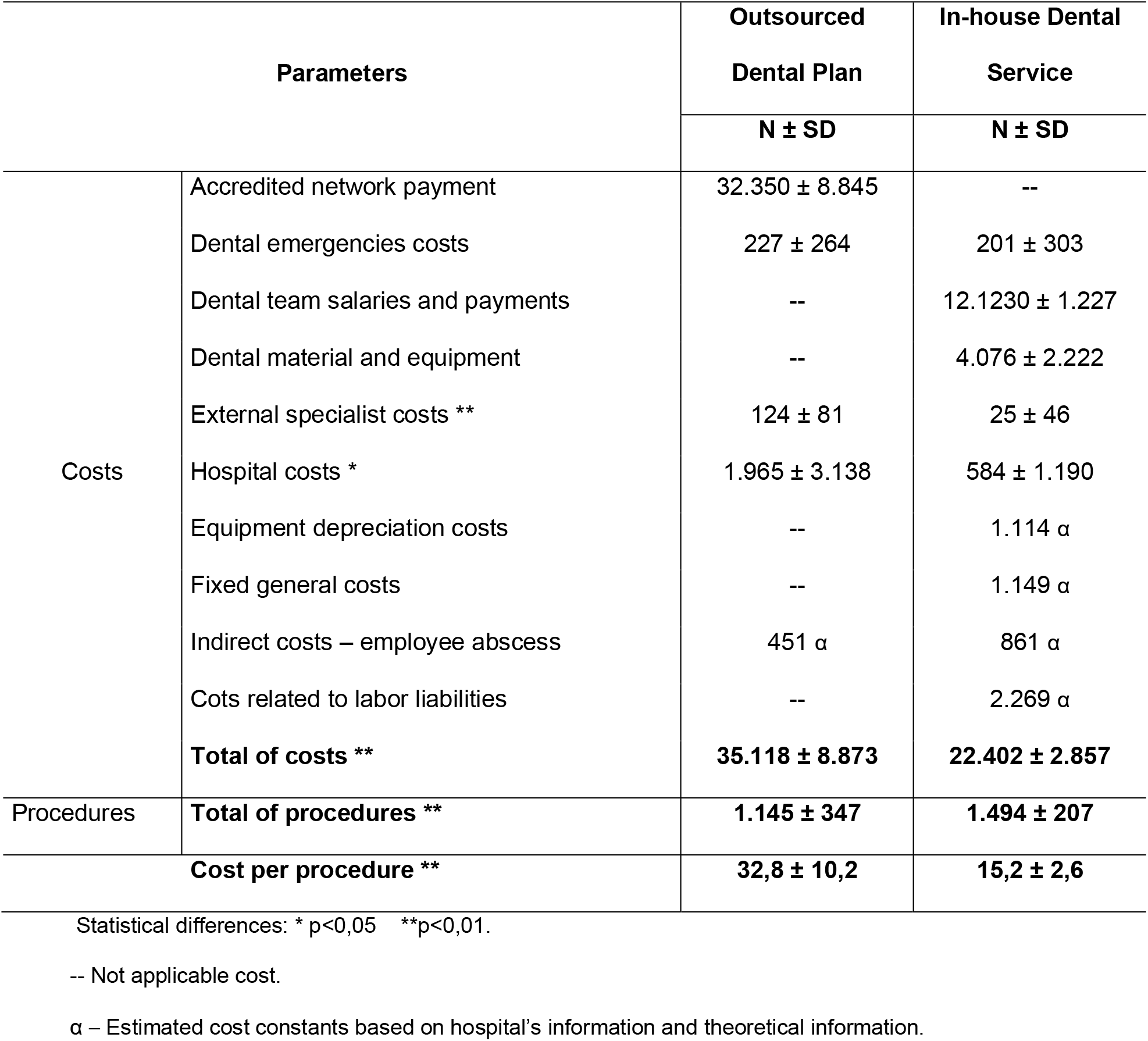
Costs (international dollars 2003) and number of procedures - Monthly means and standard deviation.

| Parameters |  | Outsourced<br>Dental Plan | In-house Dental<br>Service |
| --- | --- | --- | --- |
|  |  | N ± SD | N ± SD |
| Costs | Accredited network payment | 32.350 ± 8.845 | -- |
|  | Dental emergencies costs | 227 ± 264 | 201 ± 303 |
|  | Dental team salaries and payments | -- | 12.1230 ± 1.227 |
|  | Dental material and equipment | -- | 4.076 ± 2.222 |
|  | External specialist costs ** | 124 ± 81 | 25 ± 46 |
|  | Hospital costs * | 1.965 ± 3.138 | 584 ± 1.190 |
|  | Equipment depreciation costs | -- | 1.114 α |
|  | Fixed general costs | -- | 1.149 α |
|  | Indirect costs – employee abscess | 451 α | 861 α |
|  | Cots related to labor liabilities | -- | 2.269 α |
|  | **Total of costs \*\*** | **35.118 ± 8.873** | **22.402 ± 2.857** |
| Procedures | **Total of procedures \*\*** | **1.145 ± 347** | **1.494 ± 207** |
| **Cost per procedure \*\*** |  | **32,8 ± 10,2** | **15,2 ± 2,6** |

Statistical differences: \* p<0,05 \*\*p<0,01.
-- Not applicable cost.
α – Estimated cost constants based on hospital's information and theoretical information.

**Figure 1:**
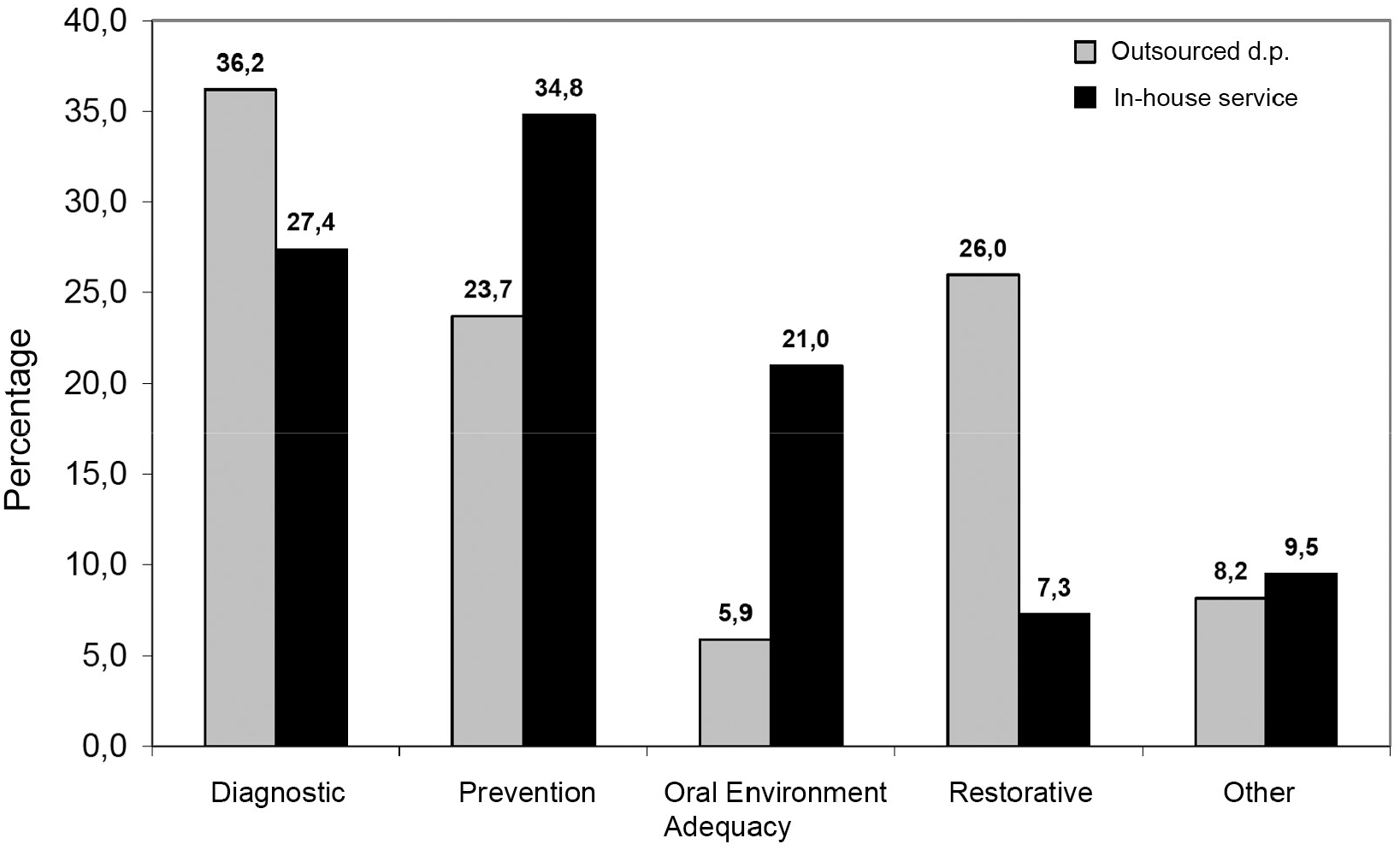
Treatment groups’ percentages at the outsourced dental plan and at the in-house dental service.

In terms of satisfaction, we found no decrease in the user satisfaction between the two services. However, this finding is based on satisfaction studies which used different methodologies and were performed by different companies.

At the beginning of service implementation, there was a fear that user satisfaction could decrease due to the restrictions on the freedom of choice of dentist. In July 2005, 83,4% of users (95% IC: between 66,5% and 93,6%) approved of the in-house service. The users’ judgments were: 30,6% excellent service; 52,8% good; 13,9% more or less satisfied; 2,8% bad service; and 0% dreadful.

Figures 2, 3, and 4 show a temporal trend of occurrence of 3 types of procedures previously mentioned in table 1 (preventive procedures, oral environment adequacy procedures and restorative procedures). These graphs show the distinct patterns of oral care between the outsourced accredited network dental plan and the in-house service.

**Figure 2:**
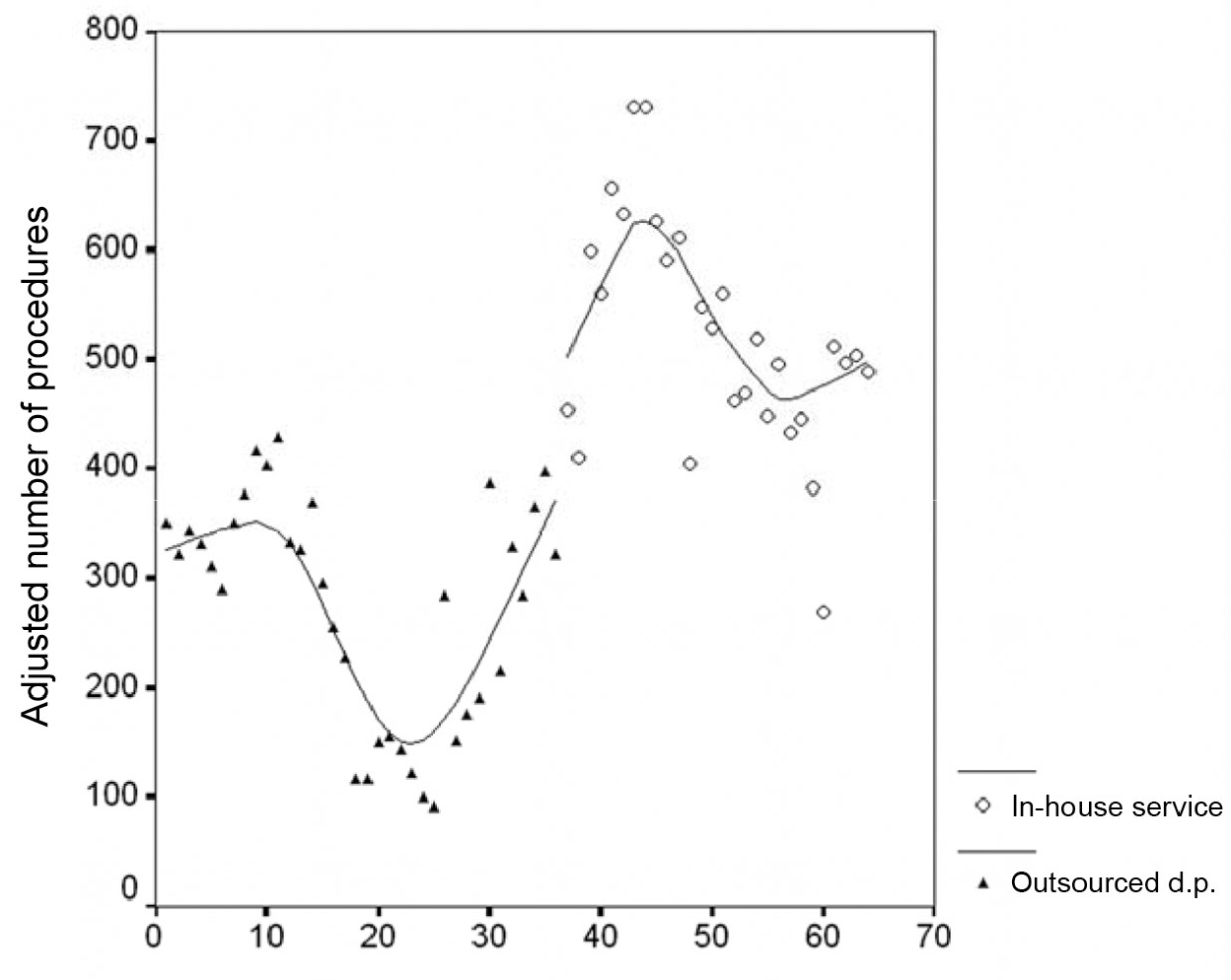
Number of preventive procedures adjusted for 4000 dental plan users – temporal trend.

**Figure 3:**
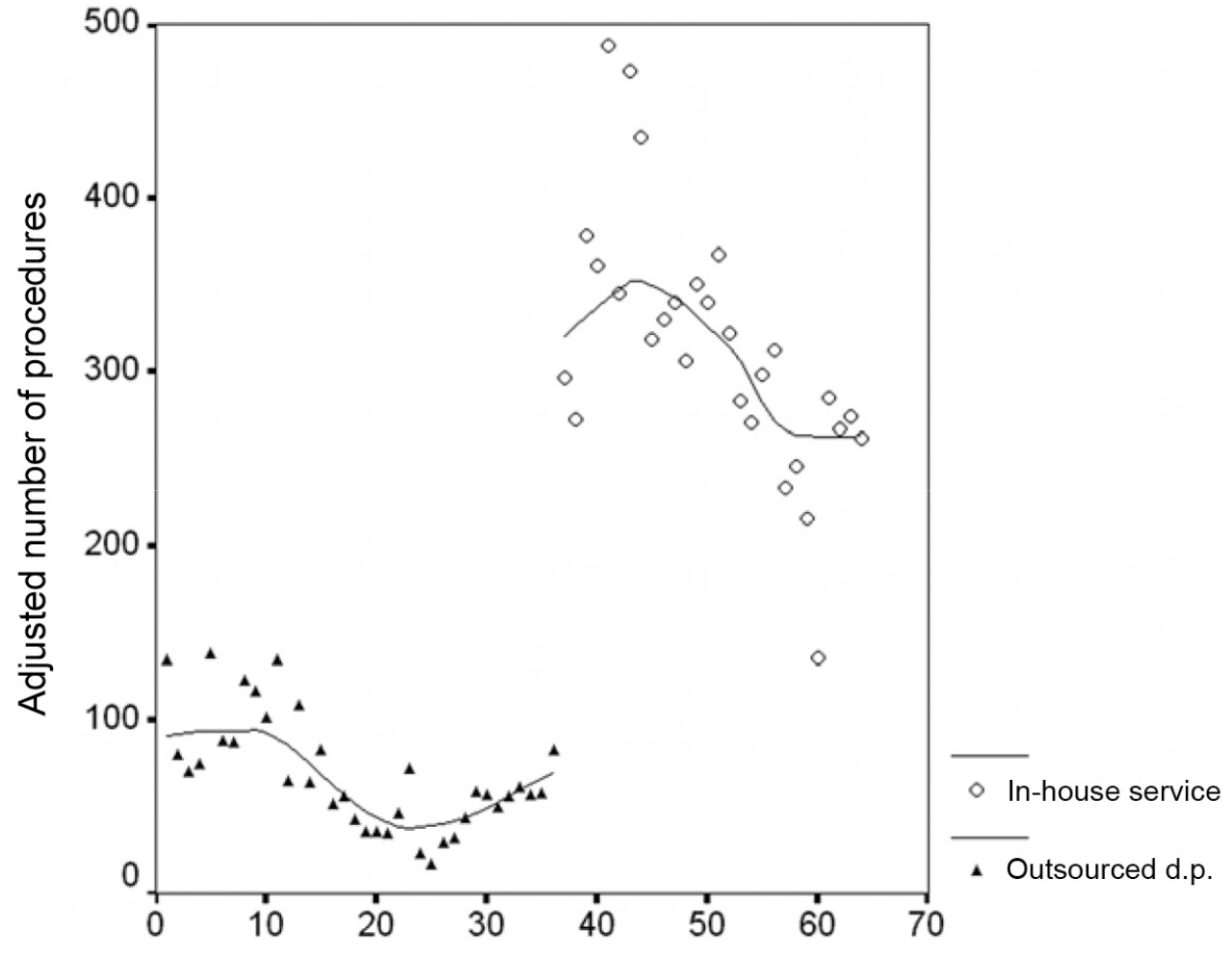
Number of oral environment adequacy procedures adjusted for 4000 dental plan users – temporal trend.

**Figure 4:**
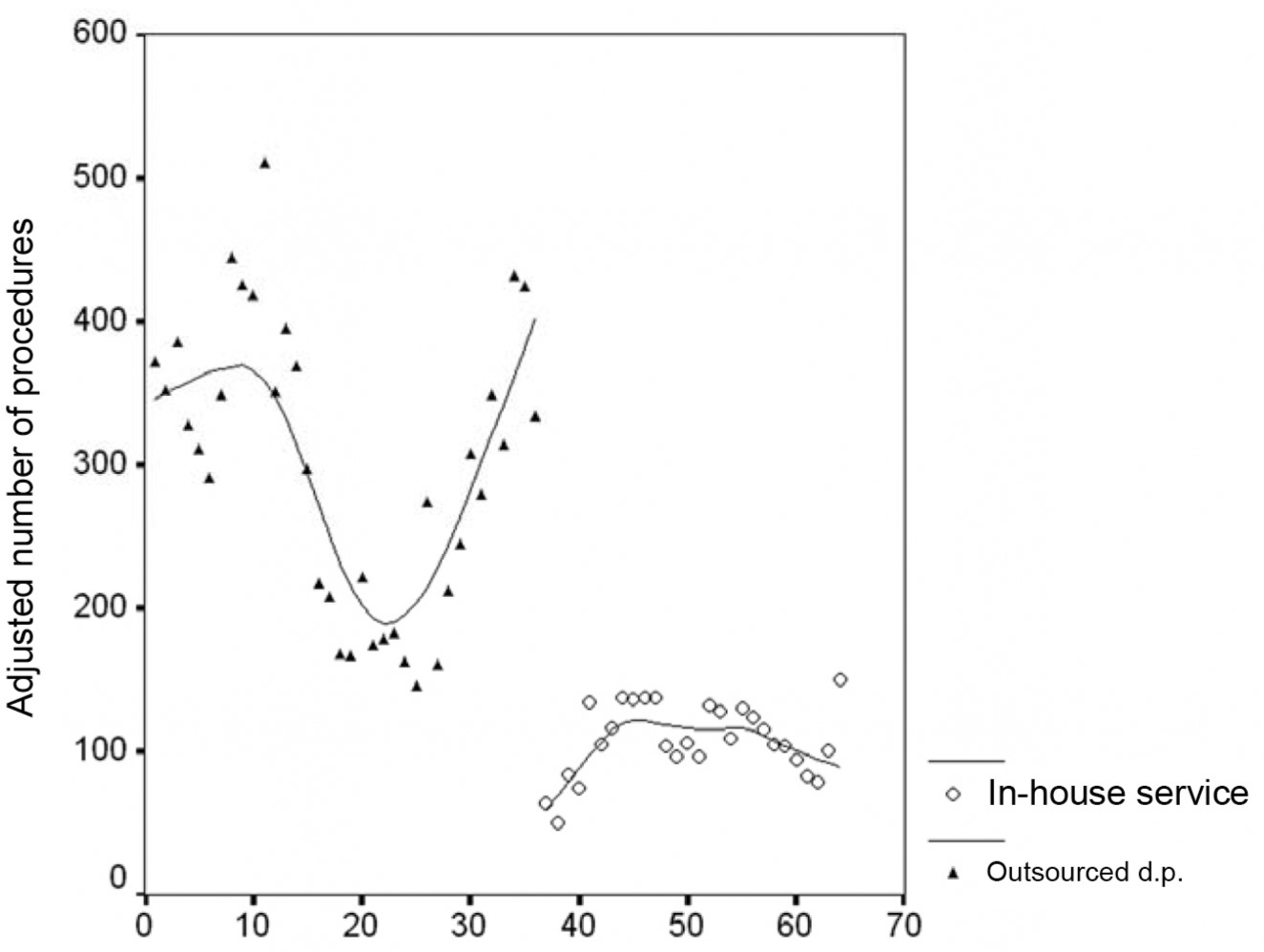
Number of restorative procedures adjusted for 4000 dental plan users – temporal trend.

Another interesting fact to observe in figures 2, 3, and 4 is that the accredited network was more susceptible to fluctuations. We could observe a decrease in the three types of procedures made by the accredited network after the 15^th^ month; this was due to a renegotiation aimed at reducing dental health plan prices. Along with the lower prices negotiated for services, a series of restrictions were placed on the dentists in the accredited network which might have caused the decline in the number of procedures done. However, this decline was followed by an increase in the number of procedures in the accredited network, which indicates that the accredited network dentists were able to learn how to bill for new procedures despite the increase in regulation. At month 36, the demand for services returned to previous levels in the accredited network, and because of this it was impossible for the outsourced dental provider to maintain the prices of the dental plan.

## DISCUSSION

Several studies have been concerned with assessing the different forms of remuneration in dentistry and their economic impact (25-30). But comparisons between health plans that operate with fee-for-service or capitation versus in-house dental services with hired dentists have not yet been evaluated. This paper presents an initial discussion of the possible effects of changing a health plan with fee-for-service payments to the dentists’ network to an in-house dental service with dentists hired as employees of the same company that provides the dental plan.

The difference in the profile of care provided by the two dental care strategies is evident, especially among the procedures that make up the greatest number of appointments, which include the areas of diagnosis, prevention, adequacy of the oral environment, and restorative procedures (table 1). These 4 groups of procedures were responsible for more than 90% of occurrences in both types of plan, but the distribution of care among them varied widely between the two oral care strategies (Figure 1).

One could argue that the decrease in restorative procedures and the increase in preventive and oral environment adequacy procedures (table 1) observed with the switch to the in-house service occurred because the demand for restorative treatments was already supplied by the outsourced dental plan. Analysis of figures 2, 3, and 4—showing the temporal occurrence of the most common types of procedures—suggests that this hypothesis does not apply because there is a sudden change of service patterns. This fact shows the difference between the two dental system’s paradigms: the in-house service is more prevention-oriented and conservative, while the accredited network is more restorative and invasive. This difference in paradigm can also be easily observed in figure 1.

Table 3 shows the creation of new areas by the in-house service: temporo-mandibular joint pathology treatments and preventive orthodontics. This was possible because before the in-house service was implemented in 2002, strong demand and desire for these areas were detected among the population. Based on this idea, evidence-based protocols were developed in order to optimize the health plan’s resources (12). This type of health management, which is concerned both with lowering health costs and with attending to employees’ needs and wishes, probably helped achieve the good quality evaluation on the customer satisfaction surveys.

Table 3 shows that the number of simple periodontal surgeries (only gingivectomy was covered under the accredited network) decreased; while the number of advanced periodontal surgeries increased (clinical crown lengthening, gingival grafts, bone regeneration surgeries, accesses flaps) and minor oral surgeries (apicectomy, removal of small cysts and lesions, etc). These results demonstrate once again that the in-house service can be even more effective in treating certain specific needs due to customization of services through a previous epidemiological study of the population’s needs. Such epidemiological surveys provided specific information and allowed the company to customize an in-house dental service with a high problem solving capability, since resources could be assigned to highly-specialized procedures for more specific but infrequent demands. Another demonstration of the in-house service’s increased capacity for problem solving in relation to the outsourced dental plan’s accredited network can be observed in table 3, which illustrates a decrease in consultations with external specialists at the in-house service and also a decrease in hospital demands, which, though infrequent, have a large impact on costs (table 4).

In table 4, it can be observed that the in-house service was able to reduce costs by 36% and yet there was an increase of over 30% in the total number of procedures, which means that the cost per procedure was reduced by more than half. One major reason for the reduction in cost was the removal of the outsourced dental plan profit component. Although the system of hired dentists is commonly accused of being less productive (27), there was no reduction in the total number of procedures with the in-house service.

Other studies comparing fee-for-service payment systems with capitation paying systems (which also have a fixed number of patients per dentist) showed that the capitation system decreases the number of procedures (30), but this fact was not observed with the in-house service analyzed in this study. Such phenomena may have happened because with capitation-based dental plans, the party which assumes the financial risk of an increase in the number of procedures is the professional (3), while with the in-house service it is the hiring company. Related to the in-house service, companies have control over the number of procedures, because the demand for procedures is proportional to the number of professionals they hire. They also have control over costs because more than half of the costs are linked to the dental staff payment (table 4), so the company is able to adjust the supply of services with a high certainty in costs. That phenomena is also observed in the general population with the entry of new professionals in the market (31).

It is important to note the methodological limitation of our study:

- Data related to the previous service were obtained through an outsourced dental plan company with an accredited network, while the data related to the in-house service were collected directly from dental records kept by professionals who were not associated with the service. This difference in data collection methods may have caused some distortions in data acquisition.
- This is not a randomized clinical trial but one company’s retrospective analysis of a health plan. Even though the population is almost the same, renewal within the employee population which could have influenced the findings. Another component of bias is that the subsequent service may have learned from the failures of the previous service.
- There are no studies comparing the effects of changing a dental plan based on an accredited network to an in-house dental service, which makes this study an isolated case that may start a new discussion. Therefore, more related studies are needed to determine the factors that influence this kind of approach in different situations and populations. It should be emphasized that the implementation of an in-house dental service without the same level of planning and management, and without inclusion of adequately skilled professionals could easily result in failure.

In low and middle-income countries few studies exist regarding economic evaluation in dentistry. The benefits of evidence-based dentistry need to be confirmed by health services research (32). In this context, the present research launches a new discussion concerning dental care cost reduction in companies as well as a paradigm shift in oral care health systems. Other cases of in-house services implementation in companies should be analyzed in order to verify the viability of this kind of assistance, as well as to discover the variables that may influence the success or failure of this kind of approach. It’s also reasonable to think that Brazil, due to its large number of dentists (1), could export experiences like the one of the present paper and also export dental teams to other countries and companies around the world.

## Data Availability

All data produced in the present study are available upon reasonable request to the authors.

http://bdtd.ibict.br/vufind/Record/URGS_079412d19c3defa510f42542d936c383

## ACKNOWLEDGMENTS

The authors would like to thank the CHOICE research group from the World Health Organization (especially Chandika Indikadahena) for sharing data information on purchasing power parity, and Drs. Ronaldo Bordin and Jandyra Fachel for their suggestions during the elaboration of this paper.

